# Structural Neuroplasticity Following Cognitive Behavioral Therapy for the Treatment of Chronic Musculoskeletal Pain: A Randomized Controlled Trial with Secondary MRI Outcomes

**DOI:** 10.1101/2021.07.13.21260466

**Authors:** James Bishop, Marina Shpaner, Antoni Kubicki, Magdalena Naylor

## Abstract

The extent of white matter (WM) and Grey matter (GM) structural neuroplasticity following cognitive behavioral therapy for chronic pain management remains undetermined. In the current study, we investigated structural alterations in GM morphometry, as well as WM complexity and connectivity, before and after an 11-week group CBT for the treatment of chronic musculoskeletal pain. We hypothesized that effective pain management would influence WM structural metrics indicative of brain plasticity, particularly within cognitive and limbic circuitry as well as GM volume within pain matrix structures. To determine this, patients were randomized into two groups: 1) CBT group that received CBT once-weekly for 11-weeks, or 2) EDU group consisting of an active patient control group that received educational materials by mail. All subjects completed behavioral assessments and underwent neuroimaging at: baseline prior to any intervention (TP1), 11-weeks following either CBT or EDU (TP2), and four months following completion of the intervention (TP3). CBT resulted in significant clinical improvements, assessed via behavioral self-reports, compared to EDU. Compared to EDU, region of interest WM analysis revealed several fiber tracts that had significantly increased WM complexity following CBT intervention, including the bilateral posterior internal capsule and the left cingulum within the temporal lobe. Conversely, several tracts exhibited a decrease in WM complexity including the right external capsule, the left posterior internal capsule, and the right cingulum within the temporal lobe. Changes in clinical outcomes were predictive of alterations in WM complexity metrics immediately following intervention and at long-term follow-up. No between-group differences were observed in either WM connectivity or GM volume. In conclusion, psychotherapeutic interventions such as group CBT influence coping strategies for effective pain relief that influence WM microstructure, however, the mechanisms of these changes remain undetermined. Future studies will be required to uncover the biological underpinnings of these alterations in pain populations.

**Clinicaltrialsgov:** Can Therapy Alter CNS Processing of Chronic Pain: A Longitudinal Study (https://clinicaltrials.gov/ct2/show/NCT01794988?term=naylor&cntry=US&state=US%3AVT&draw=2&rank=1;NCT01794988). The study protocol was registered in the Clinical Trials Database.

## Introduction

Due to high prevalence of chronic pain in society, evaluating the plasticity underlying effective treatment strategies for pain management is important for optimizing and developing novel interventions. Presently, common approaches for pain management include surgery, pharmacotherapy, and integrative health interventions, such as acupuncture and psychotherapy [18; 19]. Surgery is indicated only for select pain subtypes, carries inherent risks, and may even exacerbate symptoms in some instances such as in failed back surgery syndrome [3]. Alternatively, analgesics mask pain symptoms, but do not resolve the underlying pathophysiology of the chronic pain. In fact, long-term opioid use for chronic pain conditions is associated with poor treatment outcomes that carry the risk of addiction [13]. For these reasons, behavioral and psychotherapeutic strategies are gaining traction as safe and effective options to treat a spectrum of chronic pain disorders, including musculoskeletal pain [22].

There are a number of non-pharmacological alternatives for managing chronic pain including, not limited to cognitive behavioral therapy (CBT), yoga, and meditation. These treatments are typically aimed at increasing physical functioning, while decreasing maladaptive cognitive and behavioral phenotypes. A review of the current literature supports the short-term effectiveness of CBT for pain management, but noted long-term relapse, likely due to decreased adherence to skill practice [55]. CBT addresses aberrant psychopathologies, such as depression, anxiety, and catastrophizing that are associated with increased pain experience and psychosocial dysfunction in chronic pain populations [11]. Fear avoidance behavior is also common amongst chronic pain populations, inhibiting social and professional interactions, triggering motor dysfunction [23; 31; 41], and frequently leading to a profound loss of physical activity. Together, the triad of behavioral, psychological, and physical disturbances facilitate a positive feedback loop potentiating disability over time [47]. CBT targets these maladaptive traits by modifying emotional, behavioral, and cognitive imbalances through goal-oriented, “problem-focused” intervention.

Treatment efficacy of CBT has traditionally been demonstrated through patient self-reports, but therapeutic success has not been quantified with more objective measures of brain plasticity. More recent investigations have begun to unravel the functional network changes following CBT in pain subtypes, such as low back pain [46] and fibromyalgia [30], and migraine [44], as well CBT’s influence on network connectivity related to repeated sensory exposure to pain [28]. Structural neuroplasticity following CBT intervention, particularly WM plasticity, is less documented and may help delineate the underlying neurocircuitry involved in therapeutic efficacy. In an initial pilot study, our group previously reported that reduced maladaptive behavioral patterns, such as catastrophizing, were associated with reductions in GM volume in the dorsolateral prefrontal cortex (DLPFC) following CBT in a mixed chronic pain cohort [45]. Given the established function of the DLPFC for cognitive control and its involvement in descending pain modulation, we hypothesize that modulation of incoming nociceptive signaling through effective CBT will influence GM morphometry as well as the WM circuitry critical for relaying information within and between networks.

Although the current literature is predominated by GM structural and functional plasticity following CBT intervention, there are few large-scale randomized-controlled studies in chronic pain cohorts. Thus, the primary objectives of this study were to confirm the clinical efficacy of CBT in a randomized-controlled trial design and to determine the WM and GM neuroplasticity following 11-week CBT for musculoskeletal pain management. In addition to our primary behavioral hypothesis that CBT intervention would improve clinical measures, our primary mechanistic hypothesis was that CBT would influence WM metrics of complexity and connectivity in regions and pathways important for cognitive, affective, and emotional pain regulation while also influencing GM morphometry within the pain matrix. Based on our most recent findings of widespread WM complexity and connectivity alterations in patients with musculoskeletal pain compared to pain-free controls [8], we conducted subsequent secondary analyses with the hypothesis that effective behavioral modulation via CBT would influence WM complexity. Ancillary to this hypothesis, we conducted an exploratory whole-brain analysis of WM connectivity. Based on a previous pilot study conducted in our lab [45], we predicted that CBT would lead to the normalization of reported structural aberrations in the pain matrix, that is: a volumetric increase in regions responsible for pain modulation, such as the prefrontal (DLPFC, VMPFC), anterior cingulate (ACC), and insular cortices, and a volumetric decrease in the primary somatosensory cortex. Together, this investigation provides novel insight into the CNS neuroplasticity underlying the efficacy of CBT intervention for the treatment of chronic musculoskeletal pain.

## Materials and Methods

### Participant Recruitment and Evaluation

The study was approved by the University of Vermont Institutional Review Board (IRB). All procedures were in compliance with the Declaration of Helsinki. Prior to imaging, participants were first clinically evaluated, in-person, by a study physician and provided informed consent. Initial clinical evaluation included collection of demographic data, comprehensive medical and mental health history, medication history and management, as well as current and typical pain levels. All patients had a primary diagnosis of chronic musculoskeletal pain, which included etiologies of low back pain, osteoarthritis, post-trauma (physical), post-surgical pain, or temporomandibular disorder. Inclusion criteria for all study participants included chronic musculoskeletal pain for at least one year in duration and a minimum average of 4 out of 10 on a numerical pain Likert scale for the past month. Exclusion criteria included standard MRI contraindications, current opioid regimens, pregnancy, malignancy and/or radiation/chemotherapy, non-musculoskeletal pain disorders such as reflex sympathetic dystrophy and neuropathic pain, psychiatric and neurological disorders including major depression, schizophrenia, bipolar disorder, stroke, and epilepsy as well as other medical conditions such as a history of traumatic brain injury, diabetes, and unmanaged hypertension.

To minimize selection bias, a study statistician randomized all participants using a minimization method into either the CBT intervention group or into an educational active control group (EDU) - matched by sex, age, and pain level (pain of 4-6 and 7-10 on an 11-point scale). CBT and EDU group randomization was imbalanced [2:1] respectively and data was collected in a parallel design per institutional ethical considerations. Allocations to group were concealed to the participants and researchers collecting data. Research staff conducting the analyses were also blind to group assignment. All patients were informed by the study physician that they could cross over to open label CBT intervention following the completion of the study free of charge.

CBT consisted of weekly 90-minute group sessions for 11-weeks. CBT intervention was designed to incorporate the following pain management strategies: alter cognitions and reduce maladaptive coping strategies such as catastrophizing, promote attention-based diversion approaches, and monitor and regulate activity patterns. A thorough description of the program is outlined in the following manuscripts by Naylor and colleagues [36; 37]. All participants were required to attend the first three CBT sessions and were excluded from subsequent analyses if they missed more than two out of the eight remaining weekly sessions. Between the end of the 11-week CBT intervention and four-month long-term follow-up, individuals in the CBT group were further randomized to receive either therapeutic interactive voice response (TIVR) relapse prevention or no TIVR. Due to the minimal sample size at long-term follow-up, TIVR was not factored into the analyses.

Alternatively, pain participants randomized to the EDU control cohort received weekly educational mailings to their home pertaining broadly to pain physiology and management. These included information regarding pain chronification, physiology, positive effects of physical activity, the importance of diet and sleep patterns, as well as stress, anxiety, and depression management. Importantly, these topics were also discussed in the CBT sessions, with the critical distinction that EDU participants did not receive any information on cognitive or behavioral coping strategies.

Participants enrolled in the study underwent three separate behavioral and MRI assessments: 1) baseline, prior to treatment randomization (TP1); 2) after 11-week CBT or EDU control intervention (TP2); and 3) at a four-month follow-up after intervention (TP3) (Figure 2). At each imaging session, participants underwent an MRI evaluation consisting of structural (T_1_-weighted and diffusion-weighted imaging) and functional (resting state and task-based) acquisitions. In this manuscript, structural acquisitions were analyzed and interpreted as well as behavioral effects in a larger sample of individuals that may not have completed all imaging sessions but did complete the intervention (Supplemental Figure 1: Consort Diagram).

### Clinical Assessment

Clinical assessments were obtained by participant self-report at each study session. All participants were instructed to think about their chronic musculoskeletal pain when answering the questionnaires. Broadly, we evaluated measures of depression, pain experience, self-efficacy, and catastrophizing. Depression was measured using the Beck Depression Inventory (BDI; [6] with the following clinical parameters: < 13 = minimal depression, 14-19 = minor depression, 20-28 = moderate depression, and > 29 = major depression. Pain and disability measures were evaluated using Total Outcomes and Pain Survey (TOPS; [40]), including the following subscales: Pain Symptoms, Total Pain Experience, Perceived Family Disability, Passive Coping, as well as SF-36 Mental and Physical Health Composites. Each subscale ranged from a 0 to 100. Pain efficacy, or perceived ability to cope with chronic pain disability, was evaluated using an adapted Chronic Pain Self-Efficacy Scale [1] with the following three subscales: Self-Efficacy for Pain Management, Self-Efficacy for Physical Function, and Self-Efficacy for Coping with Symptoms. These attributes are measured on a 0-10 scale with 0 indicating “very uncertain” and 10 indicating “very certain.” Pain catastrophizing is well documented in chronic pain conditions [39] and was evaluated using the Pain Catastrophizing Scale [49]. Scores closer to 0 reflect less catastrophizing, and the maximum score of 52 is indicative of the greatest level of pain catastrophizing.

Clinical assessments from TP1, TP2, and TP3 were analyzed using SPSS Statistical software version 24.0 (IBM Corporation, Armonk, NY, USA) in the entire participant population (N=94; CBT n=60, EDU n=34). Longitudinal analyses were conducted using a repeated measures ANOVA with one within- and one between-subject factor to investigate the interaction of time (pre vs. post) and group (CBT vs. EDU).

### Magnetic Resonance Imaging Acquisition

All data were collected using a Philips 3T Achieva MRI scanner (Philips Healthcare, Best, Netherlands). The scanner underwent a hardware upgrade during the course of the study and an 8-channel sensitivity encoding (SENSE) head coil was replaced with a 15-channel SENSE digital head coil. All scan visits were completed either before or after the upgrade and data quality was assessed by a dedicated center MR physicist. The same pulse sequences were used before and after the upgrade: T_1_-weighted structural imaging data were acquired using a spoiled gradient volumetric sequence oriented perpendicular to the anterior commissure-posterior commissure with 9.9ms TR, 4.6ms TE, 8° flip angle, 256mm FOV, 1.0 NSA, 256 × 256 matrix, and 1.0mm slice thickness with no gap for 140 contiguous slices as well as an axial T2-weighted gradient spin echo sequence using 28 contiguous 5mm slices, 2466ms TR, 80ms TE, 3.0 NSA, and 230mm FOV. Diffusion weighted imaging (DWI) data were acquired using an axial 2-dimensional spin echo echo planar imaging (EPI) sequence with 46 diffusion directions and the following specifications: 59 slices, 2mm slice thickness, 10,000ms TR, 68ms TE, 2×2mm in-plane resolution, 1000 s/mm^2^ B-value, and 63 EPI factor.

### White Matter (WM) Analysis

#### 1. WM Complexity: Ball & Sticks (F1 & F2)

Two separate analyses were carried out: 1) TP1 vs. TP2, and 2) TP1 vs TP3. In the first comparison, 42 chronic pain patients receiving CBT intervention were compared to 21 active pain controls (TP1 vs. TP2). In the second analysis, 38 chronic pain patients receiving CBT intervention were compared to 15 EDU controls (TP1 vs. TP3).

WM complexity analysis was carried out as described previously [8]. Refer to Figure 1 for schematic of complexity method. In Brief, raw DWI images were first converted to nifti format using MRIcron software [42]. DWI scans were subsequently preprocessed using FSL FMRIB software [24; 48; 56]. Images underwent eddy current correction, a T_2_-weighted image was extracted from the initial B0 volume, and brain extraction (bet) was carried out on the T_2_-weighted image to create a binary mask for subsequent diffusion tensor model fitting. Single tensor (DTI) processing using the FSL dtifit tool [5] was implemented to generate FA images required for downstream analyses (described below). A multi-compartment, Ball-and-Sticks (BAS) model was implemented using the Bayesian Estimation of Diffusion Parameters Obtained Using Sampling Techniques (BEDPOSTX) to enable modeling of multiple fiber populations and orientations within each voxel [7]. Default BEDPOSTX parameters were used: a multiexponential model with a burn-in period of 1000 and a maximum of two fiber populations. The BEDPOSTX output generated estimates of each fiber orientation (i.e., x1 or x2) and a corresponding partial volume fraction metric - F1 and F2. Importantly, the model prevents overfitting by shrinking the corresponding F2 metric to 0 in the absence of a secondary fiber orientation or population. Both F1 and F2 correspond to distinct tracts within each voxel where the major or primary fiber population is defined by F1 and minor or secondary fiber population is expressed as F2.

**Figure 1.**
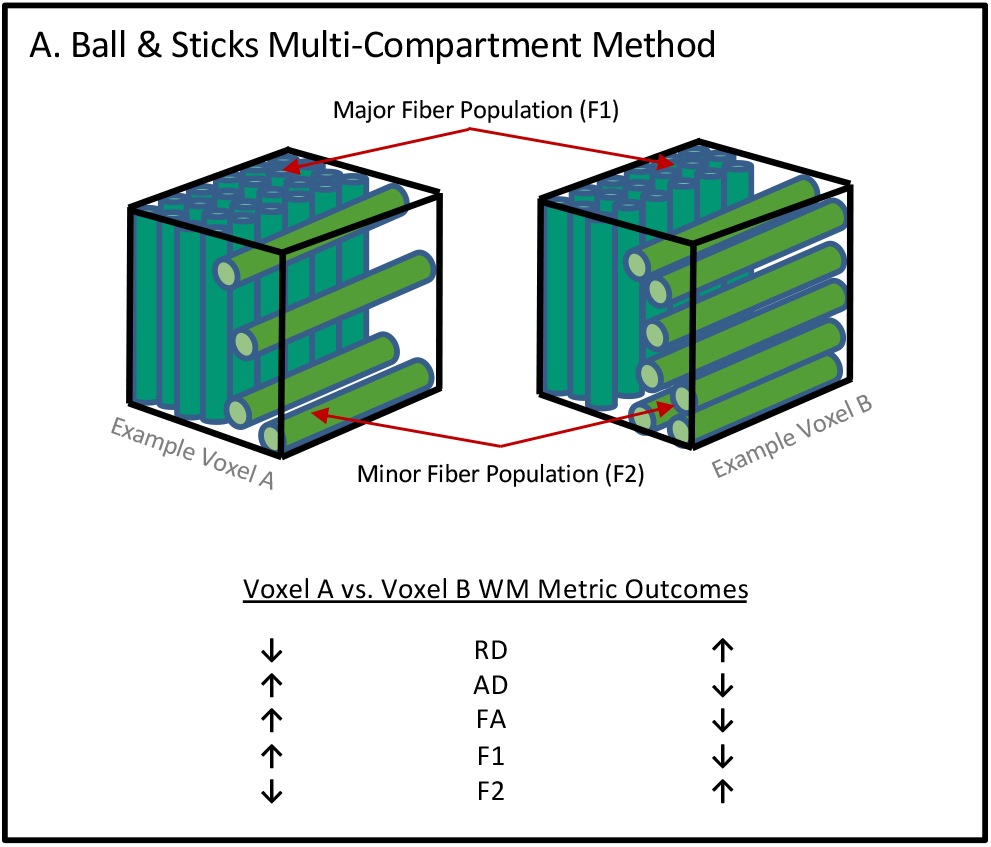
Schematic of WM Complexity. The ball & sticks multi-compartment method was utilized to define WM complexity. The “sticks” are defined by the primary (F1) and secondary (F2; if present) WM fiber populations within each voxel, after accounting for isotropic (ball) diffusion compartment. For illustrative purposes two example voxels are shown. Voxel A has a greater primary or major fiber population (F1) than voxel B and fewer secondary or minor fibers (F2). The table illustrates how partial volume fraction metrics (F1 and F2) influence commonly reported single tensor diffusion metrics (FA, RD, and AD) for each example voxel. Furthermore, this example highlights that in the case of crossing fibers, standard tensor metrics may be mistakenly interpreted as indicative of reduced fiber integrity, whereas partial volume fraction metrics will accurately separate crossing and complex fiber orientations. Figure adapted from previous manuscript [8].

Prior to inclusion, all raw diffusion scans, and metric maps were visually inspected for artifacts, excessive motion, and anatomical abnormalities resulting in the removal of four subjects from the CBT group and six subjects from the EDU group. The subsequent WM microstructural analyses steps were first carried out on the single tensor metric (FA) images using the standard FSL tract based spatial statistics (tbss) pipeline. Statistical comparison was implemented using only the partial volume maps (F1 & F2) due to their sensitivity in regions of multiple fiber populations and complex geometry. In brief, FA images were non-linearly registered to the 1×1×1mm^3^ isotropic FMRIB58 FA template followed by affine transformation to MNI152 standard space. FA data were then projected onto a mean FA skeleton image with the recommended threshold of 0.2. F1 and F2 maps were then aligned to ensure that each fiber orientation corresponded to the same fiber populations across subjects. Next, the warps generated from the non-linear and affine transformations of the FA images were applied to the F1 and F2 data. Skeletonized images for F1 and F2 were then split, and either TP2 (analysis #1) or TP3 (analysis #2) was subtracted from the TP1 baseline scan, generating a single 4D file with one difference image per subject. Statistical inference was carried out using a binary mask of the FA skeletonized image ensuring that only voxels within the center of each WM tract were analyzed, preventing partial voluming effects.

Region of interest (ROI) and whole brain exploratory analyses were conducted using FSL’s randomise method: a non-parametric, permutation approach with threshold free cluster enhancement (tfce) to correct for multiple comparisons. Whole brain voxel-by-voxel analysis was performed for each WM metric (F1 and F2) using the complete thresholded FA skeleton mask and executing a total of ten thousand permutations per voxel t-test. For the ROI analyses, masks for each of the predetermined WM structures were extracted from the Johns Hopkins ICBM-DTI-81 atlas, combined with FA skeleton mask, thresholded by a value of two to obtain the overlap between the skeleton and the mask, and binarized. Randomise permutation testing with ten thousand permutations per t-test, was then applied to each skeletonized ROI mask. Participant ages were input into the general linear model as nuisance covariates and significance was defined by P ≤ 0.05.

#### 2. Structural Connectivity Analysis

In the first comparison of baseline (TP1) to post-intervention (TP2), forty-two chronic pain patients receiving CBT intervention (CBT Group) were compared to eighteen active pain controls (EDU Group). In the second comparison of baseline (TP1) to follow-up (TP3), thirty-eight chronic pain patients receiving CBT intervention were compared to fifteen active pain controls. Structural connectivity was calculated using DSI Studio software (http://dsi-studio.labsolver.org). DWI acquisitions were reconstructed using deterministic Q-Space Dieffomorphic Reconstruction (QSDR) [59]. QSDR constructs the spin density function in template space and is a generalized form of GQI (Generalized Q-Space Imaging). Following reconstruction, whole brain tractography was initiated similarly to Muraskin and colleagues [34]. In brief, a Random number generator was created to toggle several arbitrary variables including quantitative anisotropy (QA), turning angle threshold, and smoothing. Tractography was performed 1,000 times iteratively using random whole brain seeding with the following fixed parameters: step size (1mm) and fiber length (Minimum = 10mm; Maximum = 400mm), and randomly toggled parameters: QA threshold (0.01-0.10), turning angle threshold (40°-80°), and smoothing (50%-80%). 250,000 streamlines were generated for each of the following tractography iterations.

Whole brain connectivity matrices were generated using the 116 parcellation Automated Anatomical Labeling (AAL) atlas [53]. To quantify the structural connection strength between regions, subsequent 116×116 connectivity matrices were created by counting the number of streamlines passing through and/or terminating within the respective atlas GM regions. Next, a single connectivity matrix was generated for each subject by averaging the 1,000 matrices using MATLAB 2015b Software (The MathWorks Inc., Natick, MA, 2000). Between-group differences in connectivity were statistically analyzed using the Network Based Statistics (NBS) Toolbox [60]. NBS is a validated statistical toolbox that is designed to attenuate issues of multiple comparison in mass univariate testing and facilitate large scale network comparisons. The method uses nonparametric statistical methods to control for family wise error rate. NBS is the relative equivalent of cluster-based statistical approaches except that NBS clusters in topological space instead of physical space. Each subject’s average connectivity matrix was input into the NBS toolbox to test between-group connectivity. A total of 5000 permutations were computed with family wise error (FWER) correction for multiple comparisons. An ANCOVA with 5000 permutations was computed utilizing the NBS test statistic and fully corrected for multiple comparisons via False Discovery Rate (FDR). NBS requires that a test statistic threshold is given, which can be arbitrary, and thus a range of suggested values between 2.7-3.1 were investigated in .2 increments. Significance threshold was defined at P ≤ 0.05.

### Grey Matter (GM) Analysis

Four subjects from the CBT group and six subjects from the EDU group were removed for scanner related issues and/or poor data quality. Chronic pain patients receiving CBT intervention (n=33) were compared to EDU controls (n=15) across all time points (TP1, TP2, and TP3), cumulatively consisting of 144 scans. Prior to inclusion, all raw T1-weighted anatomical scans, were visually inspected for artifacts and anatomical abnormalities. Voxel-based morphometry (VBM) analysis was implemented in SPM12 (Statistical Parametric Mapping, Institute of Neurology, London, UK) and the computational anatomy toolbox (CAT12; C. Gaser, Structural Brain Mapping Group, Jena University Hospital, Jena, Germany), using default settings. In brief, T1-weighted images were first bias-corrected for magnetic field inhomogeneities. In the CAT12 toolbox, bias-corrected images were longitudinally segmented, using the diffeomorphic anatomical registration with an exponentiated lie algebra (DARTEL) algorithm [2], into cerebrospinal fluid (CSF), grey matter (GM), and white matter (WM) compartments. The images then underwent spatial normalization to 1.5×1.5×1.5mm^3^ MNI space. Finally, all images were smoothed with an 8mm (FWHM) spatial kernel. No modulation was applied. Segmentations for each participant were visually inspected for irregularities, and sample homogeneity was assessed for outliers.

Statistical analysis was implemented using the Multivariate Repeated Measures (MRM) toolbox designed for flexible repeated measures investigation of longitudinal neuroimaging data (http://www.click2go.umip.com/i/software/mrm.html; [33]. A non-parametric, 3 (time) x 2 (groups) repeated measures MANOVA with 5000 permutations was implemented with age, sex, and upgrade as nuisance regressors and family wise error correction for multiple comparisons. Significance was assessed with a cluster threshold of P ≤ 0.05.

### Relationship Between Clinical Outcomes and WM Complexity

Post-hoc secondary analyses to determine whether changes in clinical outcome measures predicted alterations in WM complexity. Simple linear regressions were performed using the clinical change scores immediately following intervention (TP2-TP1) and at long-term follow-up (TP3-TP1) and the delta in WM complexity immediately following intervention (TP2-TP1) and long-term follow-up (TP3-TP1) using GraphPad Prism Version 8.4 (GraphPad Software, La Jolla, CA, www.graphpad.com).

## Results

### Demographics and Clinical Response to Intervention

The CBT and EDU groups were matched at the time of enrollment for age and sex, depression level, and pain duration. At immediate follow-up the groups remained balanced, however, there was a significant group difference in pain duration between study participants in the EDU control group at the long-term follow-up (TP3; Table 1). The CBT group showed significant clinical improvements over time compared to the EDU cohort on 7 out of 10 composite measures following 11-week intervention (TP2) and 4 out of 10 composite measures after four-month follow-up (TP3; Table 2).

**Table 1.** Study demographic and clinical information. Top) TP2 imaging sample demographics and clinical information indicating that the groups were balanced for age, sex, depression index, and pain duration. Bottom) TP3 imaging sample demographics indicating that the groups were balanced for age, sex, and depression, however, there was a significant between-group difference in pain duration with CBT participants having a longer pain duration than EDU controls.

| **Mean (SD)** | **CBT** | **EDU** | **P value** |
| --- | --- | --- | --- |
| Age | 46 (12.4) | 43 (12.5) | 0.37 |
| Females/Males | 34/8 | 16/5 | 0.75 |
| Depression from BDI | 16.4(8.1) | 14.2(9.5) | 0.34 |
| Pain duration | 11.7(7.9) | 8.7(8.7) | 0.18 |

Long-term Follow-up – TP3 (n (CBT) = 38, n (EDU) = 15)
| **Mean (SD)** | **CBT** | **EDU** | **P value** |
| --- | --- | --- | --- |
| Age | 46 (11.9) | 43 (12.8) | 0.42 |
| Females/Males | 31/7 | 13/2 | 1.00 |
| Depression from BDI | 16.7 (8.4) | 13.7(10.4) | 0.28 |
| Pain duration | 11.6(7.6) | 5.9(4.6) | 0.009 |

**Table 2.** Clinical improvements after 11-week intervention (TP2) and long-term follow-up (TP3). Table 2 illustrates the clinical measures collected in the trial. Immediately following 11-week intervention, CBT participants demonstrated significant clinical improvements compared to EDU controls across the majority of clinical measures including: mental pain symptoms (TOPS), physical composite score (SF-36), mental composite score (SF-36), perceived family disability (TOPS), passive coping (TOPS), catastrophizing, self-efficacy for pain management (PSE), and self-efficacy for coping with symptoms (PSE). At the long-term follow-up CBT individuals still demonstrated significant clinical improvement across several measures including passive coping (TOPS), catastrophizing, self-efficacy for pain management (PSE), and self-efficacy for coping with symptoms (CSE), however, there is evidence of potential relapse of some clinical measures. Values reported as mean ± standard deviation, CBT n=60, EDU n=34.

| Clinical Measure (SD) | CBT |  |  | EDU |  |  | TP1 vs. TP2 (CBT vs. EDU) |  | TP1 vs. TP2 vs. TP3 (CBT vs. EDU) |  |
| --- | --- | --- | --- | --- | --- | --- | --- | --- | --- | --- |
|  | TP1 | TP2 | TP3 | TP1 | TP2 | TP3 | TimeXGroup F | TimeXGroup P | TimeXGroup F | TimeXGroup P |
| Total Pain Experience (TOPS) | 51.00 (14.28) | 39.49 (15.11) | 36.02 (14.53) | 53.27 (15.84) | 46.10 (16.26) | 41.90 (17.34) | 3.751 | 0.056 | 1.795 | 0.172 |
| Pain Symptoms (TOPS) | 63.93 (13.78) | 51.71 (19.78) | 47.45 (18.00) | 65.37 (13.09) | 59.57 (17.66) | 55.6 (18.70) | 4.311 | 0.041 | 2.101 | 0.129 |
| Physical Composite Score (SF-36) | 31.94 (9.10) | 38.27 (9.27) | 39.53 (9.97) | 31.14 (8.50) | 34.45 (9.08) | 36.91 (10.26) | 4.812 | 0.031 | 2.123 | 0.126 |
| Mental Composite Score (SF-36) | 43.78 (12.37) | 49.34 (11.12) | 48.35 (11.64) | 44.31 (13.94) | 44.42 (13.75) | 45.79 (11.02) | 4.546 | 0.036 | 2.924 | 0.059 |
| Perceived Family Disability (TOPS) | 45.64 (20.8) | 33.22 (22.27) | 27.24 (19.16) | 47.5 (22.73) | 38.07 (21.78) | 33.20 (20.69) | 0.609 | 0.437 | 0.492 | 0.613 |
| Passive Coping (TOPS) | 38.53 (19.56) | 31.15 (19.53) | 25.65 (18.97) | 34.93 (24.96) | 35.30 (19.80) | 33.93 (19.46) | 5.022 | 0.027 | 5.139 | 0.008 |
| Catastrophizing | 19.91 (10.5) | 9.95 (7.56) | 8.16 (7.8) | 21.2 (10.8) | 17.43 (10.58) | 16.37 (12.27) | 10.029 | 0.002 | 5.306 | 0.007 |
| Self-Efficacy for Pain Management (PSE) | 49.42 (18.54) | 67.56 (20.57) | 68.87 (21.05) | 46.07 (16.43) | 50.53 (20.69) | 57.60 (22.79) | 11.21 | 0.001 | 5.646 | 0.005 |
| Self-Efficacy for Physical Function (FSE) | 72.42 (16.86) | 81.49 (17.11) | 82.25 (16.02) | 67.30 (26.96) | 73.07 (24.23) | 76.96 (23.07) | 2.268 | 0.132 | 1.346 | 0.266 |
| Self-Efficacy for Coping with Symptoms (CSE) | 52.14 (16.68) | 70.68 (15.94) | 71.20 (16.36) | 49.67 (16.03) | 52.63 (20.56) | 58.00 (20.60) | 31.1 | <0.001 | 13.759 | <0.001 |

### WM Region of Interest Complexity Analyses: (Baseline (TP1) vs. Post-Intervention (TP2))

A subtraction (TP2 – TP1) analysis assessing baseline and post 11-week intervention visits was implemented to compare EDU controls to patients receiving CBT with age, sex, and scanner upgrade as nuisance covariates. The ball & sticks multi-compartment metric of fiber complexity, F1, yielded several significant between-group differences at a threshold of p<0.05. Specifically, the CBT cohort exhibited an increase in fiber complexity (a reduction in the major fiber population F1), compared to EDU controls, within the left posterior internal capsule and the right external capsule (Table 3). In addition, the CBT group exhibited greater fiber complexity (increased F2), compared to EDU controls, within the left cingulum adjacent to the hippocampus, the bilateral posterior internal capsule. The left posterior internal capsule, therefore, exhibited increased CBT-related complexity, as revealed by both, the major and the minor fiber population metrics.

**Table 3.** ROI complexity results after 11-week intervention (TP2). At TP2, CBT participants exhibited an increase in complexity compared to EDU, bilaterally in the cingulum adjacent to the hippocampus, right external capsule, and left posterior internal capsule. EDU Controls demonstrated an increase in complexity compared to CBT participants within the right posterior internal capsule. Significance was evaluated at using threshold free cluster enhancement (TFCE) with a threshold of p ≤ 0.05, CBT n=42, EDU n=21.

| WM ROI | Hemisphere | F1 |  | F2 |  |
| --- | --- | --- | --- | --- | --- |
|  |  | *CBT > EDU* | *CBT < EDU* | *CBT > EDU* | *CBT < EDU* |
| Anterior Internal Capsule | Left |  |  |  |  |
|  | Right |  |  |  |  |
| Cerebral Peduncle | Left |  |  |  |  |
|  | Right |  |  |  |  |
| Cingulum | Left |  |  |  |  |
|  | Right |  |  |  |  |
| Cingulum Adjacent to Hippocampus | Left |  |  | ✓ |  |
|  | Right |  |  |  |  |
| External Capsule | Left |  |  |  |  |
|  | Right |  | ✓ |  |  |
| Posterior Internal Capsule | Left |  | ✓ | ✓ |  |
|  | Right |  |  | ✓ |  |
| Uncinate Fasciculus | Left |  |  |  |  |
|  | Right |  |  |  |  |

### WM Region of Interest Complexity Analyses: Baseline (TP1) vs. Final Follow-up (TP3)

CBT group exhibited increased fiber complexity (increased F2) when we compared CBT vs. EDU from baseline to final follow-up. Significant increases in F2 were observed within left posterior internal capsule and the right anterior internal capsule. Increased complexity in the left posterior internal capsule was, therefore, consistent across the entire study (Table 4).

**Table 4.** ROI complexity results after intervention follow-up (TP3). At TP3, CBT participants exhibited an increase in complexity compared to EDU in the right anterior internal capsule and the posterior internal capsule. EDU Controls demonstrated an increase in complexity compared to CBT participants within the right cerebral peduncle. Significance was evaluated using threshold free cluster enhancement (TFCE) with a threshold of p ≤ 0.05, CBT n=38, EDU n=15.

| WM ROI | Hemisphere | F1 |  | F2 |  |
| --- | --- | --- | --- | --- | --- |
|  |  | *CBT > EDU* | *CBT < EDU* | *CBT > EDU* | *CBT < EDU* |
| Anterior Internal Capsule | Left |  |  |  |  |
|  | Right |  |  | ✓ |  |
| Cerebral Peduncle | Left |  |  |  |  |
|  | Right |  |  |  |  |
| Cingulum | Left |  |  |  |  |
|  | Right |  |  |  |  |
| Cingulum Adjacent to Hippocampus | Left |  |  |  |  |
|  | Right |  |  |  |  |
| External Capsule | Left |  |  |  |  |
|  | Right |  |  |  |  |
| Posterior Internal Capsule | Left |  | ✓ | ✓ |  |
|  | Right |  |  |  |  |
| Uncinate Fasciculus | Left |  |  |  |  |
|  | Right |  |  |  |  |

### WM Whole Brain Complexity Analyses: Baseline (TP1) vs. Post-Follow-up (TP2)

A subtraction (TP2 - TP1) analysis assessing baseline and post-intervention visits was implemented to compare EDU controls to patients receiving CBT with age, sex, and scanner upgrade as nuisance covariates. Whole brain permutation analyses of the ball & sticks multi-compartment metric, F1, did not yield any significant between-group results at a threshold of p<.05. The ball & sticks multi-compartment metric, F2, also did not yield any significant between-group results at a threshold of p<.05, however several notable clusters were observed. These include increased F2, or the minor fiber population metric, in the left anterior corona radiata (p=0.08) and the left genu of the corpus callosum (p=0.1) of participants receiving CBT.

### WM Whole Brain Complexity Analyses: Pre- (Tp1) vs. Follow-up (Tp3)

An analysis assessing baseline and four-month post-intervention follow-up visits was implemented to compare EDU controls to patients receiving CBT with age, sex, and scanner upgrade as nuisance covariates. Whole brain permutation analyses of both ball & sticks multi-compartment metrics, F1 and F2, did not yield any significant between-group results at a threshold of p<.05.

### WM Connectivity

A longitudinal comparison of connectivity did not yield any significant whole brain differences between patients receiving CBT compared to EDU controls (Table 5A). A significant effect of time was observed, with both groups demonstrating less connectivity at TP3 compared to TP1 evaluation at t-value thresholds of 2.9 (p<0.036) and 3.1 (p<0.041) (Table 5B)

**Table 5.** Longitudinal whole brain WM connectivity. A) Longitudinal analysis of connectivity corrected for multiple comparisons demonstrated a main effect of time at threshold values 2.9 and 3.1, with both CBT and EDU participants exhibiting reduced connectivity at TP3 compared to TP1. This finding was not significant at a threshold of 2.7. No significant group by time effects in connectivity were observed following TP2 or TP3 study visits. Significance was evaluated at threshold p ≤ 0.05, CBT n=38, EDU n=12. B) Increased connectivity was observed between cortical and subcortical regions several t-value thresholds.

| Longitudinal (TP1, TP2, TP3) - CBT= 38; EDU=12 |  |  |  |
| --- | --- | --- | --- |
| Contrast | t-value Threshold | Permutations | Lowest p-value |
| TP1>TP2 | 3.1 | 5000 | 0.94 |
| TP1>TP3 | 3.1 | 5000 | 0.04 |
| TP1<TP2 | 3.1 | 5000 | 0.66 |
| TP1<TP3 | 3.1 | 5000 | 0.66 |
| CBT>EDU (TP1>TP2) | 3.1 | 5000 | 0.44 |
| CBT>EDU (TP1>TP3) | 3.1 | 5000 | 0.12 |
| EDU>CBT (TP1>TP2) | 3.1 | 5000 | 0.94 |
| EDU>CBT (TP1>TP3) | 3.1 | 5000 | 0.94 |
| TP1>TP2 | 2.9 | 5000 | 0.87 |
| TP1>TP3 | 2.9 | 5000 | 0.03 |
| TP1<TP2 | 2.9 | 5000 | 0.70 |
| TP1<TP3 | 2.9 | 5000 | 0.87 |
| CBT>EDU (TP1>TP2) | 2.9 | 5000 | 0.69 |
| CBT>EDU (TP1>TP3) | 2.9 | 5000 | 0.20 |
| EDU>CBT (TP1>TP2) | 2.9 | 5000 | 0.99 |
| EDU>CBT (TP1>TP3) | 2.9 | 5000 | 0.68 |
| TP1>TP2 | 2.7 | 5000 | 0.31 |
| TP1>TP3 | 2.7 | 5000 | 0.07 |
| TP1<TP2 | 2.7 | 5000 | 0.91 |
| TP1<TP3 | 2.7 | 5000 | 0.81 |
| CBT>EDU (TP1>TP2) | 2.7 | 5000 | 0.55 |
| CBT>EDU (TP1>TP3) | 2.7 | 5000 | 0.31 |
| EDU>CBT (TP1>TP2) | 2.7 | 5000 | 0.89 |
| EDU>CBT (TP1>TP3) | 2.7 | 5000 | 0.51 |

\*No Covariates

B.
| TP1>TP3 |  |  |  |
| --- | --- | --- | --- |
| Connection | Test Statistic | t-value Thresh. | Significance (p-value) |
| Hippocampus R. to Lingual L. | 3.60 | 3.1 | 0.041 |
| Cingulum Post. R. to Lingual L. | 3.44 | 3.1 | 0.041 |
| Amygdala R. to Lingual L. | 3.13 | 3.1 | 0.041 |
| Connection | Test Statistic | t-value Thresh. | Significance (p-value) |
| Hippocampus R. to Lingual L. | 3.60 | 2.9 | 0.036 |
| Cingulum Post. R. to Lingual L. | 3.44 | 2.9 | 0.036 |
| Amygdala R. to Lingual L. | 3.13 | 2.9 | 0.036 |
| Frontal Med. Orb. R. to Temporal Sup. R. | 3.05 | 2.9 | 0.036 |
| Frontal Med. Orb. R. to Heschl R. | 2.94 | 2.9 | 0.036 |
| Frontal Med. Orb. R. to Supramarginal R. | 2.91 | 2.9 | 0.036 |

### GM Whole Brain Morphometry Analyses (Longitudinal – TP1, TP2, & TP3)

Longitudinal analyses assessing baseline (TP1), post 11-week intervention (TP2), and follow-up (TP3) visits were implemented using the Multivariate and Repeated Measures for Neuroimaging (MRM) toolbox. To investigate the possible main effects of group and group by time interactions, permutation testing was conducted with age, sex, and upgrade as nuisance regressors, family-wise error correction for multiple comparisons, and a cluster threshold of P ≤ 0.05. No significant main effects or interactions were observed at the predetermined statistical threshold of P ≤ 0.05.

### Clinical Outcomes Predict WM Complexity

Changes in several WM complexity metrics were significantly predicted by changes clinical outcomes across timepoints. Immediate post (TP2-TP1): Self-efficacy for coping with symptoms (CSE) predicted alterations in WM minor fiber populations (F2) within the right posterior internal capsule *R*^*2*^ = .129, *F*_(1,60)_ = 8.944, *p* = .004 and the left posterior internal capsule *R*^*2*^ = .077, *F*_(1,60)_ = 5.040, *p* = .028. Changes in CSE immediately following intervention also predicted changes in WM major fiber population (F1) in the right external capsule *R*^*2*^ = .114, *F*_(1,60)_ = 7.773, *p* = .007. A composite mean self-efficacy score which consists of CSE, self-efficacy for physical function (FSE), and self-efficacy for pain management (PSE) also predicted alterations in the WM minor fiber population (F2) within the right cingulum adjacent to the hippocampus immediately following intervention *R*^*2*^ = .096, *F*_(1,60)_ = 6.401, *p* = .014 (Figure 3). Long-term follow-up (TP3-TP1): Total pain experience (TOPS) was predictive of WM alterations in the left posterior internal capsule for both major (F1) *R*^*2*^ = .087, *F*_(1,50)_ = 4.818, *p* = .032 and minor (F2) *R*^*2*^ = .110, *F*_(1,50)_ = 6.185, *p* = .016 fiber populations. Similar to immediate post, CSE also predicted changes in WM minor fiber population (F2) within the left posterior internal capsule at long-term follow-up *R*^*2*^ = .126, *F*_(1,49)_ = 7.119, *p* = .01. Additionally, the change in composite mean self-efficacy score (CSE, PSE, FSE) was predictive of change in WM minor fiber population (F2) within the left posterior internal capsule *R*^*2*^ = .099, *F*_(1,50)_ = 5.508, *p* = .022 (Figure 4).

**Figure 2.**
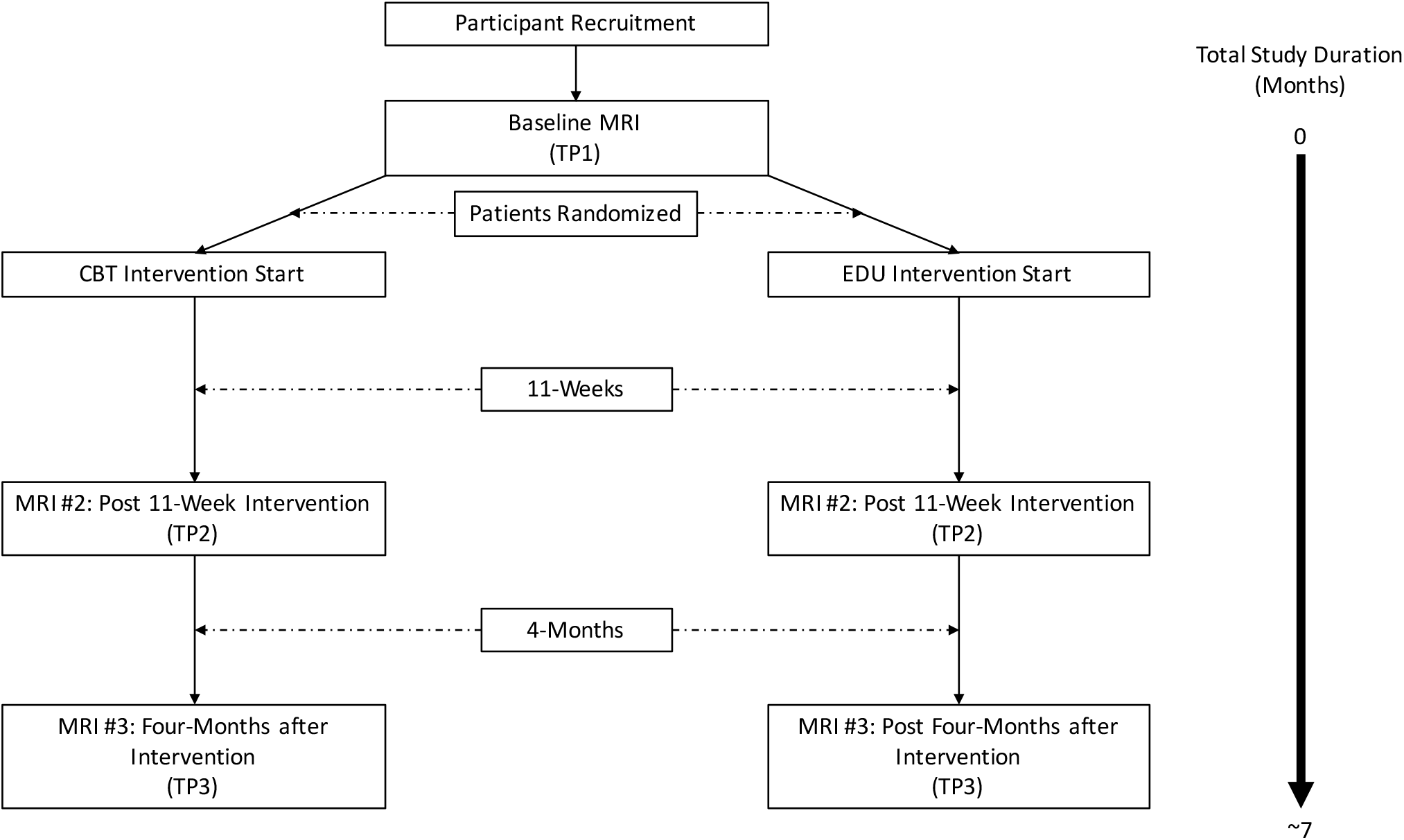
Study Design. Study flow chart illustrating the total study enrollment duration and the data collection timeline. All chronic pain patients underwent a baseline (TP1) MRI session and were subsequently randomized to either CBT intervention or an active educational materials control group (EDU). Both CBT and EDU interventions were 11-weeks in duration after which all participants underwent a second, identical MRI session (TP2). Patients were then brought back for a third MRI session four months after completion of either CBT or EDU intervention (TP3). Total study duration from time of initial visit to completion of the final MRI was approximately seven months for each participant.

**Figure 3.**
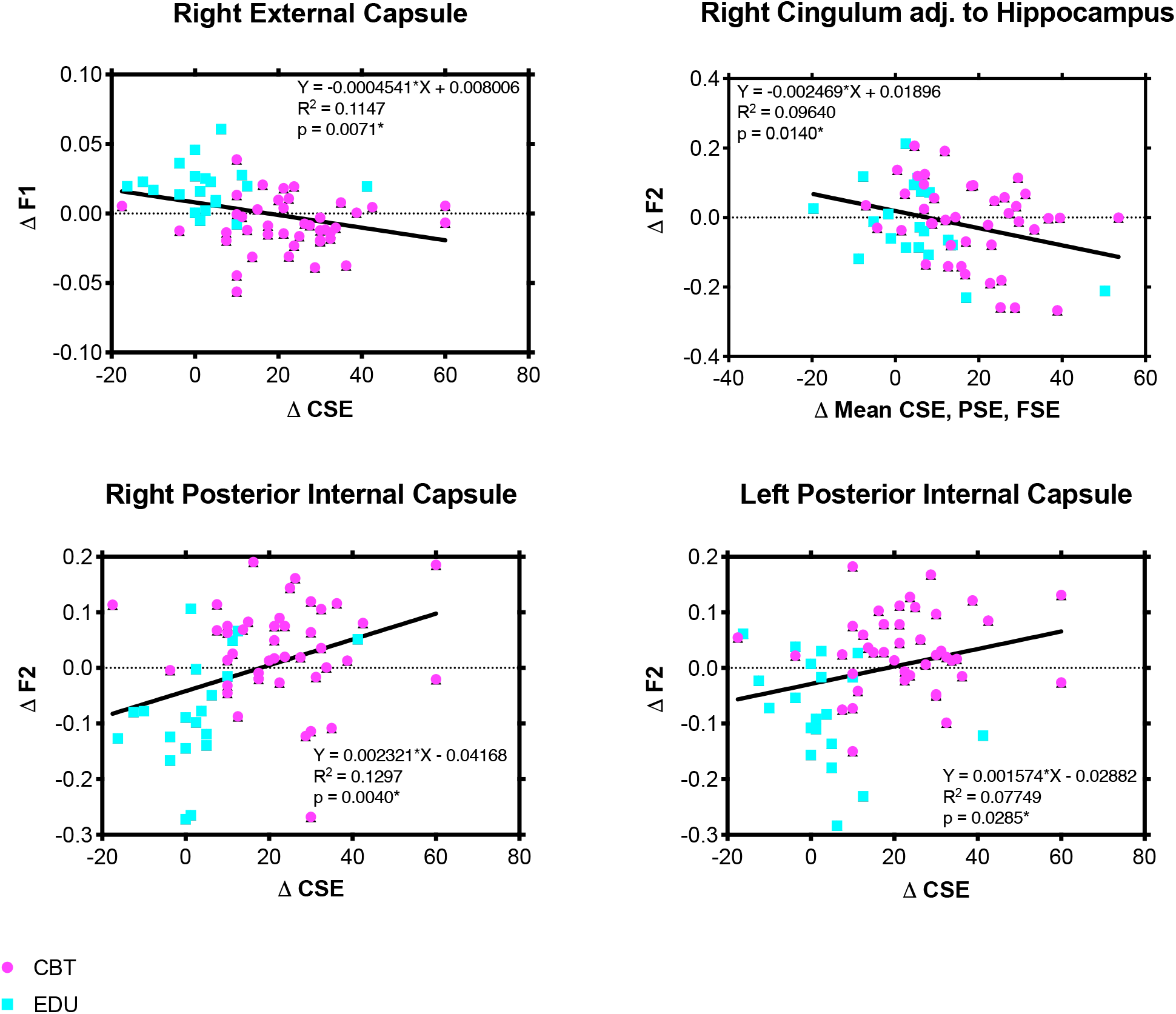
Relationship Between Changes in WM Complexity Metrics and Clinical Self-Reports Immediately Following Intervention (TP2 vs TP1) Across Musculoskeletal Pain Participants. WM values were extracted from tract clusters demonstrating between-group changes in complexity immediately following 11-week intervention (TP2 vs. TP1). Linear regression plots illustrating the relationship between the change in WM complexity metrics (F1 and F2) and the change in clinical self-efficacy behavioral measures (self-efficacy for coping with symptoms = CSE; self=efficacy for pain management = PSE; and self-efficacy for physical functioning = FSE) at 11-weeks from baseline assessment. Colors indicate the study group for each participant (CBT=purple; EDU=Blue), however, the relationship was examined across all participants without factoring in group assignment.

**Figure 4.**
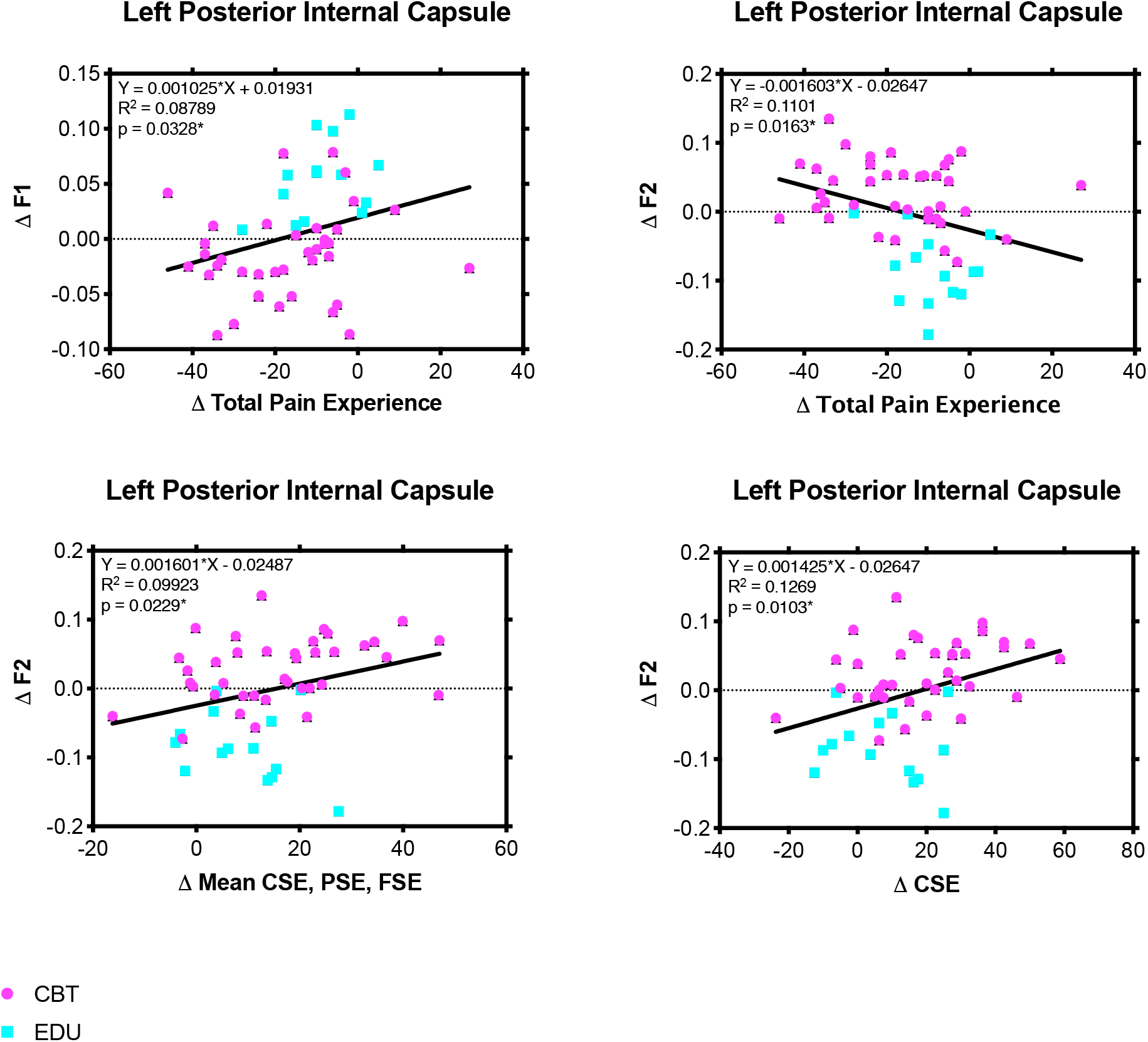
Relationship Between Changes in WM Complexity Metrics and Clinical Self-Reports at 1-Month Post Intervention Follow-up (TP3 vs TP1) Across Musculoskeletal Pain Participants. WM values were extracted from tract clusters demonstrating between-group changes in complexity immediately following long-term follow-up (TP3 vs. TP1). Linear regression plots illustrating the relationship between the change in WM complexity metrics (F1 and F2) and the change in clinical behavioral measures (total pain experience = TOPS; self-efficacy for coping with symptoms = CSE; self=efficacy for pain management = PSE; and self-efficacy for physical functioning = FSE) at long-term follow-up from baseline assessment. Colors indicate the study group for each participant (CBT=purple; EDU=Blue), however, the relationship was examined across all participants without factoring in group assignment.

## Discussion

In this investigation, we longitudinally evaluated structural neuroplasticity via metrics of WM complexity, WM connectivity, and GM morphometry in chronic musculoskeletal pain patients randomized to either CBT intervention or an active educational control group (EDU). To our knowledge, this is the first randomized clinical study in a chronic pain population to report CBT-related WM neuroplasticity. Building on our previous report demonstrating baseline differences between chronic musculoskeletal pain and healthy pain-free controls [8], we demonstrated significant between-group alterations in WM fiber complexity immediately following 11-week intervention (TP2) and at six-month long-term follow-up (TP3). Post-hoc exploratory regression analyses collapsed across groups demonstrated significant relationships between clinical improvements across a spectrum of motor, sensory and cognitive outcome measures and WM structural alterations of complexity suggesting either compensation or normalization of aberrant behavior may factor into neuroplasticity irrespective of intervention type. Although it is challenging to directly link microstructural mechanisms with the utilized outcome measures, central sensitization in chronic musculoskeletal pain contributes to by dysfunctional signaling in the brain and spinal cord [43; 61] and thus is subject to activity-dependent neuroplasticity [16]. Interventions designed to influence aberrant behaviors such as CBT provide a window for therapeutic efficacy that, based on our findings, are capable of influencing WM metrics of complexity in the adult brain. This activity-dependent neuroplasticity concept has been shown focally using electro-acupuncture where WM changes have been demonstrated in the somatosensory cortex following clinical improvements in median nerve latency [32]. Thus, we demonstrate that learning to cope with chronic pain through CBT intervention facilitates dynamic changes in WM microstructure across sensorimotor, affective, and cognitive networks.

### Sensorimotor Pathways

Sensorimotor tracts, including the posterior limb of the internal capsule, involve pathways through which sensory and motor signals are relayed between the brain and spinal cord, and are central in developing chronic pain. The posterior internal capsule is comprised of multiple fiber bundles that influence nociception and associated behavioral alterations. These include ascending fibers that project to the somatosensory cortex, descending motor fiber projections, as well as thalmo-cortical afferent and efferent projections [14]. Following intervention, CBT patients exhibited a bilateral increase in posterior internal capsule complexity, however, only the left posterior internal capsule remained significantly more complex after the final follow-up. Because the internal capsule contains multiple fiber pathways it is challenging to dissect the specific modality of the CBT related effects. Post hoc analyses comparing the relationship between motor and sensory behavioral measures such as composite self-efficacy scores and total pain experience with WM metrics further corroborates multimodal effects in the left posterior internal capsule. Neurosurgical reports using electrical stimulation applied to the posterior internal capsule have been shown to reduce pain in patients with thalamic pain syndrome posited to be facilitated by engagement of the descending pain-modulation networks via cortical-thalamic connections [14]. WM infrastructure is required to propagate transmission of sensory information and thus neuroplastic changes may underlie CBT efficacy especially via increased cognitive control of pain-modulation networks. This concept of influencing white matter by modulation of pain signaling has been shown focally in carpal tunnel syndrome following acupuncture [32]. Across CBT and EDU control groups, an increase in the minor fiber population (F2) was identified and changes were predicted by composite self-efficacy measures. Increases in self-efficacy following CBT for pain management have been reported [25; 27; 46] and also linked to extinction of fear [62]. Maladaptive fear avoidance behavior in chronic pain populations contributes to pain chronification by limiting a patient’s physical functioning and further potentiating aberrations in peripheral structure and pain signaling [29]. Our findings indicate overall improvements in composite self-efficacy scores, however, between-group physical functioning self-efficacy subscales were not significant following intervention at either timepoint alluding to a more prominent role of the sensory networks.

### Emotion Processing Pathways

Affective dysregulation is central to the development of chronic pain [58] as well as subsequent CNS alterations [21]. Our results suggest that CBT aimed at reversing maladaptive traits by accomplishing better affective regulation for pain management are reflected in plasticity of the neural tracts that integrate affective information. This is supported by clinical improvements in affective pain behaviors (self-efficacy, coping, etc.) as well as significant between-group differences within the external capsule immediately post-therapy (TP2) – a tract that revealed structural alterations at baseline compared to pain-free healthy controls in our previous report [8]. The external capsule contains projections to the amygdala and the insular cortex, important nodes of the affective-motivational aspects of pain. Pain participants receiving CBT showed a reduction in the primary fiber population metric, F1, compared to EDU pain controls in the external capsule which were predicted by a change in self-efficacy for coping with symptoms immediately following intervention. Interestingly, the F1 metric within the external capsule was reduced at baseline in pain patients compared to pain-free healthy control in our previous cross-sectional investigation[8]. A further reduction in this WM metric following CBT may be indicative of a compensatory plasticity or alternatively influencing discrete fiber populations within the WM bundle. External capsule differences between active and control intervention groups were not maintained at long-term follow-up (TP3) which is consistent with behavioral and symptom relapse identified in several clinical measures. Potential relapse is a recognized challenge for CBT with as many as 30-70% of individuals relapsing after initial improvements [52]. We have previously established remote tools for relapse prevention [37] which require further study to evaluate whether neuroplasticity can be either maintained or increased with continued adherence and relapse mitigation.

The cingulum bundle in the temporal lobe adjacent to the hippocampus demonstrated increased WM fiber complexity following CBT intervention (TP2), which was absent at follow-up (TP3). Compared to EDU, CBT participants demonstrated an increase in F2 (minor fiber population) with the left hemisphere. The cingulum provides connections between an array of brain regions in the temporal, parietal, and frontal lobes [57]. The temporal lobe subcomponent of the cingulum is particularly relevant to pain processing and memory encoding given projections to the hippocampus, the entorhinal cortex, and the cingulate cortex, another node in the affective-motivational pain network. In this context, surgical lesions to the cingulate and cingulum have been performed as a treatment for numerous intractable chronic pain states, including low back pain and have been shown to diminish the affective response to noxious stimuli (See Reviews: [9; 15]). WM plasticity cingulum immediately following intervention was predicted by composite self-efficacy scores encompassing pain, coping, and physical functioning further supporting changes in the affective appraisal of chronic pain following CBT.

### Cognitive Pathways

We also found increased CBT-related complexity in the anterior limb of the internal capsule at TP3. This WM bundle is important for cognitive processing of pain because it connects the thalamus to the prefrontal cortex, including the DLPFC [17]. Alterations in the thalamo-cortical pathway at TP3 in CBT patients may be indicative of long-term changes in cognitive processing of pain. Engagement of the DLPFC provides several therapeutic alternatives for chronic pain including engagement of the endogenous opioid system which has been demonstrated using neuromodulation approaches such as transcranial magnetic stimulation [51]. Although we also hypothesized that CBT would influence DLPFC GM morphometry similar to what we previously reported [45], no significant between group-differences were identified in the current study. This null finding is consistent with a recent randomized clinical trial implementing mindfulness-based stress reduction for mitigation of migraine frequency [44].

### Caveats

There are several important limitations that warrant consideration:

1. <u>Heterogeneity of Chronic Musculoskeletal Pain Type:</u> Pain is a highly subjective experience that can be influenced by biological, social, and cultural factors. Compounding these factors are co-morbidities, such as anxiety and depression, that have increased prevalence in pain populations. Studies from around the world reveal that up to 64% of patients, who experience chronic pain, have multiple sources of pain [10; 20; 35; 50; 54]. Different chronic pain etiologies may differentially influence brain structure [4]; however, involvement of higher order pain processing regions is consistently shown [38] and the multidimensional aspect of pain suggests complex mechanisms [12] - even within disorders. In this manuscript, we report structural neuroplasticity common across multiple musculoskeletal subtypes. Interventions focusing on a single type of chronic musculoskeletal pain have limited feasibility and real-world significance.
2. <u>Potential Performance Bias:</u> Performance bias is difficult to control in the context of a behavioral intervention. It is possible that participants could figure out from available online resources that the educational materials intervention serves as an active control despite the fact that we never explicitly disclosed this. Importantly, baseline imaging was conducted prior to disclosure of the randomization assignment. Characterization of expectation effects on neuroplasticity is an interesting avenue for future research.
3. <u>Temporal Effects of Plasticity:</u> The biological and physiological mechanisms underlying observed WM alterations in neuroimaging metrics are not well-understood, and the precise temporal evolution of these changes may vary depending on many factors such as the specificity or duration of an intervention. For example, Maeda and colleagues demonstrated changes in anisotropy of the WM adjacent to the motor cortex following electro-acupuncture-induced improvements in median nerve latency for carpal tunnel syndrome [32]. These WM changes were observed in a similar timeframe (3-months) and demonstrated somatotopic specificity. Heterogeneity in individual treatment adherence and therapeutic strategy choice may dilute the detectible WM changes in a 3-month timeframe. Long-term investigations of WM, combined with objective readouts of practice adherence and skill use can uncover reliable targets of plasticity.
4. <u>MRI Upgrade:</u> The influence of scanner upgrade on brain WM metrics is important to consider. The imaging protocol is not expected to have influenced the contrast or distortion of the collected images. There is a possibility of introducing miniscule bias due to increased signal-to-noise ratio (SNR) following the upgrade. The testing conducted by the dedicated center physicists did not yield any significant effect of upgrade when evaluating the noise floor for the diffusion weighted imaging acquisitions. The literature also supports the stability of within-scanner measurements, following upgrades [26].

## Conclusions

In conclusion, CBT provides a promising non-pharmacological treatment alternative to reduce symptoms of musculoskeletal pain. Our results suggest that 11-week CBT is capable of promoting WM neuroplasticity across sensorimotor, affective, and cognitive fiber pathways. We propose that WM plasticity reflects alterations in aberrant pain signaling through effective remediation of maladaptive behavioral traits that are reminiscent of the chronic pain condition. However, additional investigations are required to confirm plasticity findings addressing some of the limitations in the current study and to define the specific neural mechanisms underlying changes in fiber complexity.

## Data Availability

Data available upon request.

## Financial Disclosures / Acknowledgments

Research reported in this publication was supported by the National Institute of Arthritis and Musculoskeletal and Skin Diseases (NIAMS), of the National Institutes of Health under award number R01AR059674 (to MN). The content is solely the responsibility of the authors and does not necessarily represent the official views of the National Institutes of Health.

The authors would like to thank the UVM MRI Center for Biomedical Imaging for their assistance with data acquisition and the Vermont Advanced Computing Core (VACC), which is supported by NASA (NNX-08AO96G), for providing high performance computing resources.

The authors (JB, MS, AK, MN) report no financial interest or potential conflicts of interest.

---

**Supplemental Figure 1.**
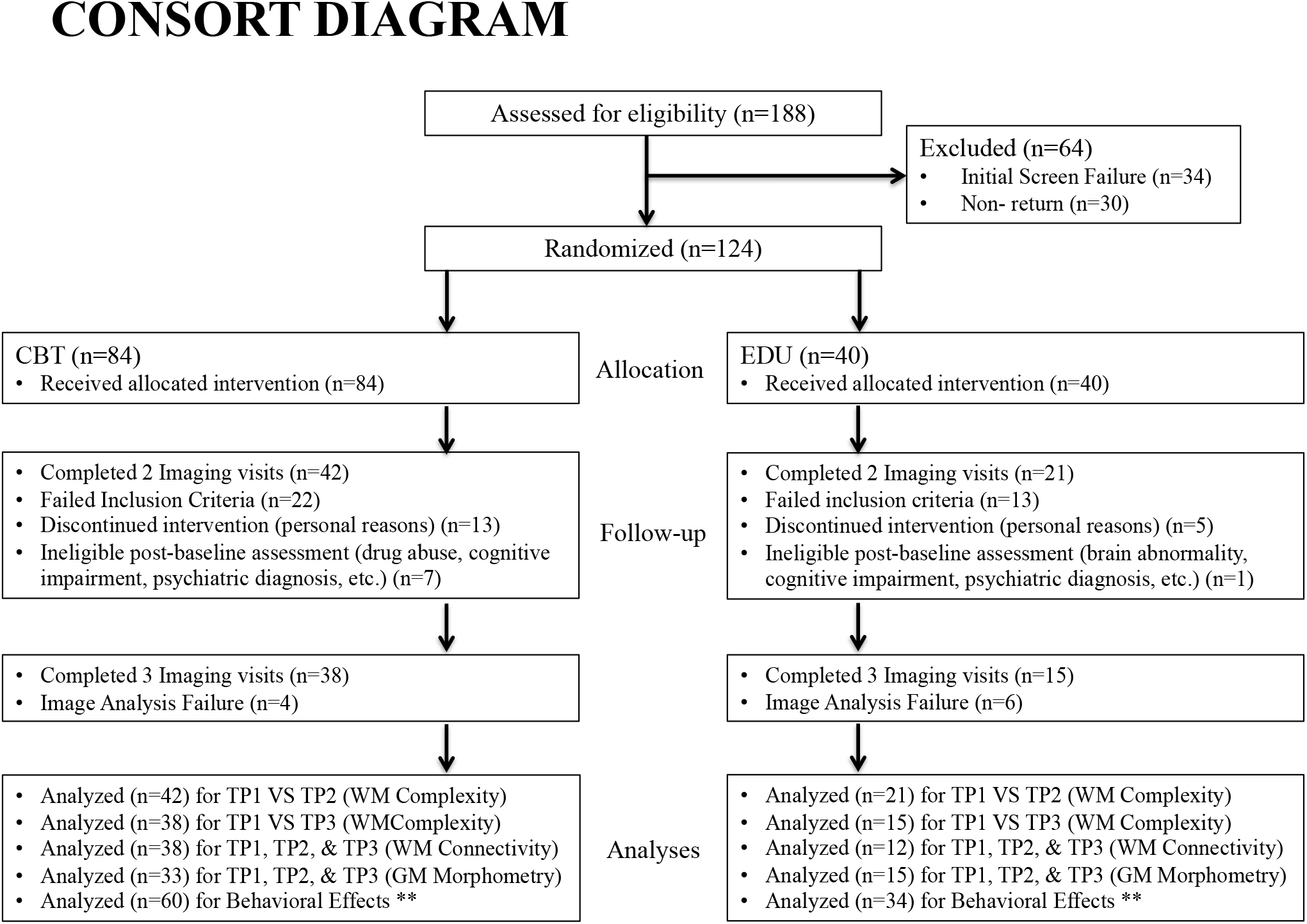
Consort Diagram.

## References

[1] Anderson KO, Dowds BN, Pelletz RE, Edwards WT, Peeters-Asdourian C. Development and initial validation of a scale to measure self-efficacy beliefs in patients with chronic pain. Pain 1995;63(1):77–84.

[2] Ashburner J. A fast diffeomorphic image registration algorithm. Neuroimage 2007;38(1):95–113.

[3] Baber Z, Erdek MA. Failed back surgery syndrome: current perspectives. J Pain Res 2016;9:979–987.

[4] Baliki MN, Schnitzer TJ, Bauer WR, Apkarian AV. Brain morphological signatures for chronic pain. PLoS One 2011;6(10):e26010.

[5] Basser PJ, Mattiello J, LeBihan D. Estimation of the effective self-diffusion tensor from the NMR spin echo. J Magn Reson B 1994;103(3):247–254.

[6] Beck AT, Ward CH, Mendelson M, Mock J, Erbaugh J. An inventory for measuring depression. Arch Gen Psychiatry 1961;4:561–571.

[7] Behrens TE, Berg HJ, Jbabdi S, Rushworth MF, Woolrich MW. Probabilistic diffusion tractography with multiple fibre orientations: What can we gain? Neuroimage 2007;34(1):144–155.

[8] Bishop JH, Shpaner M, Kubicki A, Clements S, Watts R, Naylor MR. Structural network differences in chronic muskuloskeletal pain: Beyond fractional anisotropy. Neuroimage 2018;182:441–455.

[9] Bubb EJ, Metzler-Baddeley C, Aggleton JP. The cingulum bundle: Anatomy, function, and dysfunction. Neurosci Biobehav Rev 2018;92:104–127.

[10] Dominick CH, Blyth FM, Nicholas MK. Unpacking the burden: understanding the relationships between chronic pain and comorbidity in the general population. Pain 2012;153(2):293–304.

[11] Ehde DM, Dillworth TM, Turner JA. Cognitive-behavioral therapy for individuals with chronic pain: efficacy, innovations, and directions for research. Am Psychol 2014;69(2):153–166.

[12] Elman I, Borsook D. Common Brain Mechanisms of Chronic Pain and Addiction. Neuron 2016;89(1):11–36.

[13] Els C, Jackson TD, Kunyk D, Lappi VG, Sonnenberg B, Hagtvedt R, Sharma S, Kolahdooz F, Straube S. Adverse events associated with medium-and long-term use of opioids for chronic non-cancer pain: an overview of Cochrane Reviews. The Cochrane database of systematic reviews 2017;10:CD012509.

[14] Franzini A, Messina G, Levi V, D’Ammando A, Cordella R, Moosa S, Prada F, Franzini A. Deep brain stimulation of the posterior limb of the internal capsule in the treatment of central poststroke neuropathic pain of the lower limb: case series with long-term follow-up and literature review. J Neurosurg 2019:1–9.

[15] Fuchs PN, Peng YB, Boyette-Davis JA, Uhelski ML. The anterior cingulate cortex and pain processing. Front Integr Neurosci 2014;8:35.

[16] Ganguly K, Poo MM. Activity-dependent neural plasticity from bench to bedside. Neuron 2013;80(3):729–741.

[17] Giguere M, Goldman-Rakic PS. Mediodorsal nucleus: areal, laminar, and tangential distribution of afferents and efferents in the frontal lobe of rhesus monkeys. J Comp Neurol 1988;277(2):195–213.

[18] Giller CA. The neurosurgical treatment of pain. Arch Neurol 2003;60(11):1537–1540.

[19] Grabois M. Management of chronic low back pain. Am J Phys Med Rehabil 2005;84(3 Suppl):S29–41.

[20] Gureje O, Von Korff M, Kola L, Demyttenaere K, He Y, Posada-Villa J, Lepine JP, Angermeyer MC, Levinson D, de Girolamo G, Iwata N, Karam A, Guimaraes Borges GL, de Graaf R, Browne MO, Stein DJ, Haro JM, Bromet EJ, Kessler RC, Alonso J. The relation between multiple pains and mental disorders: results from the World Mental Health Surveys. Pain 2008;135(1-2):82–91.

[21] Hashmi JA, Baliki MN, Huang L, Baria AT, Torbey S, Hermann KM, Schnitzer TJ, Apkarian AV. Shape shifting pain: chronification of back pain shifts brain representation from nociceptive to emotional circuits. Brain 2013;136(Pt 9):2751–2768.

[22] Hassett AL, Williams DA. Non-pharmacological treatment of chronic widespread musculoskeletal pain. Best Pract Res Clin Rheumatol 2011;25(2):299–309.

[23] Hodges PW, Tucker K. Moving differently in pain: a new theory to explain the adaptation to pain. Pain 2011;152(3 Suppl):S90–98.

[24] Jenkinson M, Beckmann CF, Behrens TE, Woolrich MW, Smith SM. Fsl. Neuroimage 2012;62(2):782–790.

[25] Jensen MP, Turner JA, Romano JM. Changes in beliefs, catastrophizing, and coping are associated with improvement in multidisciplinary pain treatment. J Consult Clin Psychol 2001;69(4):655–662.

[26] Jovicich J, Czanner S, Han X, Salat D, van der Kouwe A, Quinn B, Pacheco J, Albert M, Killiany R, Blacker D, Maguire P, Rosas D, Makris N, Gollub R, Dale A, Dickerson BC, Fischl B. MRI-derived measurements of human subcortical, ventricular and intracranial brain volumes: Reliability effects of scan sessions, acquisition sequences, data analyses, scanner upgrade, scanner vendors and field strengths. Neuroimage 2009;46(1):177–192.

[27] Kashikar-Zuck S, Sil S, Lynch-Jordan AM, Ting TV, Peugh J, Schikler KN, Hashkes PJ, Arnold LM, Passo M, Richards-Mauze MM, Powers SW, Lovell DJ. Changes in pain coping, catastrophizing, and coping efficacy after cognitive-behavioral therapy in children and adolescents with juvenile fibromyalgia. The journal of pain : official journal of the American Pain Society 2013;14(5):492–501.

[28] Kucyi A, Salomons TV, Davis KD. Cognitive behavioral training reverses the effect of pain exposure on brain network activity. Pain 2016;157(9):1895–1904.

[29] Langevin HM, Sherman KJ. Pathophysiological model for chronic low back pain integrating connective tissue and nervous system mechanisms. Med Hypotheses 2007;68(1):74–80.

[30] Lazaridou A, Kim J, Cahalan CM, Loggia ML, Franceschelli O, Berna C, Schur P, Napadow V, Edwards RR. Effects of Cognitive-Behavioral Therapy (CBT) on Brain Connectivity Supporting Catastrophizing in Fibromyalgia. Clin J Pain 2017;33(3):215–221.

[31] Lund JP, Donga R, Widmer CG, Stohler CS. The pain-adaptation model: a discussion of the relationship between chronic musculoskeletal pain and motor activity. Can J Physiol Pharmacol 1991;69(5):683–694.

[32] Maeda Y, Kim H, Kettner N, Kim J, Cina S, Malatesta C, Gerber J, McManus C, Ong-Sutherland R, Mezzacappa P, Libby A, Mawla I, Morse LR, Kaptchuk TJ, Audette J, Napadow V. Rewiring the primary somatosensory cortex in carpal tunnel syndrome with acupuncture. Brain 2017;140(4):914–927.

[33] McFarquhar M, McKie S, Emsley R, Suckling J, Elliott R, Williams S. Multivariate and repeated measures (MRM): A new toolbox for dependent and multimodal group-level neuroimaging data. Neuroimage 2016;132:373–389.

[34] Muraskin J, Dodhia S, Lieberman G, Garcia JO, Verstynen T, Vettel JM, Sherwin J, Sajda P. Brain dynamics of post-task resting state are influenced by expertise: Insights from baseball players. Hum Brain Mapp 2016;37(12):4454–4471.

[35] Murphy KR, Han JL, Yang S, Hussaini SM, Elsamadicy AA, Parente B, Xie J, Pagadala P, Lad SP. Prevalence of Specific Types of Pain Diagnoses in a Sample of United States Adults. Pain Physician 2017;20(2):E257–E268.

[36] Naylor MR, Helzer JE, Naud S, Keefe FJ. Automated telephone as an adjunct for the treatment of chronic pain: a pilot study. The journal of pain : official journal of the American Pain Society 2002;3(6):429–438.

[37] Naylor MR, Keefe FJ, Brigidi B, Naud S, Helzer JE. Therapeutic Interactive Voice Response for chronic pain reduction and relapse prevention. Pain 2008;134(3):335–345.

[38] Ong WY, Stohler CS, Herr DR. Role of the Prefrontal Cortex in Pain Processing. Mol Neurobiol 2019;56(2):1137–1166.

[39] Quartana PJ, Campbell CM, Edwards RR. Pain catastrophizing: a critical review. Expert Rev Neurother 2009;9(5):745–758.

[40] Rogers WH, Wittink HM, Ashburn MA, Cynn D, Carr DB. Using the “TOPS,” an outcomes instrument for multidisciplinary outpatient pain treatment. Pain Med 2000;1(1):55–67.

[41] Roland MO. A critical review of the evidence for a pain-spasm-pain cycle in spinal disorders. Clin Biomech (Bristol, Avon) 1986;1(2):102–109.

[42] Rorden C, Brett M. Stereotaxic display of brain lesions. Behav Neurol 2000;12(4):191–200.

[43] Roussel NA, Nijs J, Meeus M, Mylius V, Fayt C, Oostendorp R. Central sensitization and altered central pain processing in chronic low back pain: fact or myth? Clin J Pain 2013;29(7):625–638.

[44] Seminowicz DA, Burrowes SAB, Kearson A, Zhang J, Krimmel SR, Samawi L, Furman AJ, Keaser ML, Gould NF, Magyari T, White L, Goloubeva O, Goyal M, Peterlin BL, Haythornthwaite JA. Enhanced mindfulness-based stress reduction in episodic migraine: a randomized clinical trial with magnetic resonance imaging outcomes. Pain 2020.

[45] Seminowicz DA, Shpaner M, Keaser ML, Krauthamer GM, Mantegna J, Dumas JA, Newhouse PA, Filippi CG, Keefe FJ, Naylor MR. Cognitive-behavioral therapy increases prefrontal cortex gray matter in patients with chronic pain. The journal of pain : official journal of the American Pain Society 2013;14(12):1573–1584.

[46] Shpaner M, Kelly C, Lieberman G, Perelman H, Davis M, Keefe FJ, Naylor MR. Unlearning chronic pain: A randomized controlled trial to investigate changes in intrinsic brain connectivity following Cognitive Behavioral Therapy. Neuroimage Clin 2014;5:365–376.

[47] Shpaner M, Tulipani LJ, Bishop JH, Naylor MRJCBNR. The Vicious Cycle of Chronic Pain in Aging Requires Multidisciplinary Non-pharmacological Approach to Treatment. 2017;4(3):176–187.

[48] Smith SM, Jenkinson M, Woolrich MW, Beckmann CF, Behrens TE, Johansen-Berg H, Bannister PR, De Luca M, Drobnjak I, Flitney DE, Niazy RK, Saunders J, Vickers J, Zhang Y, De Stefano N, Brady JM, Matthews PM. Advances in functional and structural MR image analysis and implementation as FSL. Neuroimage 2004;23 Suppl 1:S208–219.

[49] Sullivan M, Bishop S, Pivak J. The pain catastrophizing scale: development and validation. Psychological Assessment 1995;7 524–532.

[50] Suri P, Morgenroth DC, Kwoh CK, Bean JF, Kalichman L, Hunter DJ. Low back pain and other musculoskeletal pain comorbidities in individuals with symptomatic osteoarthritis of the knee: data from the osteoarthritis initiative. Arthritis Care Res (Hoboken) 2010;62(12):1715–1723.

[51] Taylor JJ, Borckardt JJ, George MS. Endogenous opioids mediate left dorsolateral prefrontal cortex rTMS-induced analgesia. Pain 2012;153(6):1219–1225.

[52] Turk DC, Rudy TE. Neglected topics in the treatment of chronic pain patients--relapse, noncompliance, and adherence enhancement. Pain 1991;44(1):5–28.

[53] Tzourio-Mazoyer N, Landeau B, Papathanassiou D, Crivello F, Etard O, Delcroix N, Mazoyer B, Joliot M. Automated anatomical labeling of activations in SPM using a macroscopic anatomical parcellation of the MNI MRI single-subject brain. Neuroimage 2002;15(1):273–289.

[54] Watkins EA, Wollan PC, Melton LJ, 3rd, Yawn BP. A population in pain: report from the Olmsted County health study. Pain Med 2008;9(2):166–174.

[55] Williams AC, Eccleston C, Morley S. Psychological therapies for the management of chronic pain (excluding headache) in adults. The Cochrane database of systematic reviews 2012;11:Cd007407.

[56] Woolrich MW, Jbabdi S, Patenaude B, Chappell M, Makni S, Behrens T, Beckmann C, Jenkinson M, Smith SM. Bayesian analysis of neuroimaging data in FSL. Neuroimage 2009;45(1 Suppl):S173–186.

[57] Wu Y, Sun D, Wang Y, Wang Y. Subcomponents and Connectivity of the Inferior Fronto-Occipital Fasciculus Revealed by Diffusion Spectrum Imaging Fiber Tracking. Front Neuroanat 2016;10:88.

[58] Yang S, Chang MC. Chronic Pain: Structural and Functional Changes in Brain Structures and Associated Negative Affective States. Int J Mol Sci 2019;20(13).

[59] Yeh FC, Tseng WY. NTU-90: a high angular resolution brain atlas constructed by q-space diffeomorphic reconstruction. Neuroimage 2011;58(1):91–99.

[60] Zalesky A, Fornito A, Bullmore ET. Network-based statistic: identifying differences in brain networks. Neuroimage 2010;53(4):1197–1207.

[61] Zhang L, Zhou L, Ren Q, Mokhtari T, Wan L, Zhou X, Hu L. Evaluating Cortical Alterations in Patients With Chronic Back Pain Using Neuroimaging Techniques: Recent Advances and Perspectives. Front Psychol 2019;10:2527.

[62] Zlomuzica A, Preusser F, Schneider S, Margraf J. Increased perceived self-efficacy facilitates the extinction of fear in healthy participants. Front Behav Neurosci 2015;9:270.

